# Vaccination and three non-pharmaceutical interventions determine the end of COVID-19 at 381 metropolitan statistical areas in the US

**DOI:** 10.1101/2021.05.18.21257362

**Authors:** Lu Zhong, Mamadou Diagne, Qi Wang, Jianxi Gao

## Abstract

The rapid rollout of the COVID-19 vaccine global raises the question of whether and when the ongoing pandemic could be eliminated with vaccination and non-pharmaceutical interventions (NPIs). Despite advances in the impact of NPIs and the conceptual belief that NPIs and vaccination control COVID-19 infections, we lack evidence to employ control theory in real-world social human dynamics in the context of disease spreading. We bridge the gap by developing a new analytical framework that treats COVID-19 as a feedback control system with the NPIs and vaccination as the controllers and a computational and mathematical model that maps human social behaviors to input signals. This approach enables us to effectively predict the epidemic spreading in 381 Metropolitan statistical areas (MSAs) in the US by learning our model parameters utilizing the time series NPIs (i.e., the stay-at-home order, face-mask wearing, and testing) data. This model allows us to optimally identify three NPIs to predict infections actually in 381 MSAs and avoid overfitting. Our numerical results universally demonstrate our approach’s excellent predictive power with *R*^2^ > 0.9 of all the MSAs regardless of their sizes, locations, and demographic status. Our methodology allows us to estimate the needed vaccine coverage and NPIs for achieving *R*_*e*_ to the manageable level and the required days for disease elimination at each location. Our analytical results provide insights into the debates on the aims for eliminating COVID-19. NPIs, if tailored to the MSAs, can drive the pandemic to an easily containable level and suppress future recurrences of epidemic cycles.

---

The ongoing global pandemic of coronavirus disease 2019 (COVID-19) has caused devastating loss of human lives and inflicted severe economic burden in the US^1^. By 20 March 2021, more than 510,000 people were killed by COVID-19, the unemployment reaches 11.5%^2^, and fiscal shortfall reaches over 200 billion^3^. In response to the disease, federal, state, and local governments have implemented non-pharmaceutical interventions (NPIs) from encouragement and recommendations to full-on regulation and sanctions^4–6^ and pharmaceutical interventions (PIs) with available vaccinations^7, 8^ and drugs^9^. However, these NPIs (e.g., social distancing, face mask-wearing, hand hygiene, testing, contact tracing, isolation, etc.) are often loosened and re-tightened without rigorous empirical evidence. Methodologies, from randomized controlled trials^5^, econometric methods^10, 11^ to mathematical models^12–18^, have been developed to measure the effects of these NPIs. But many of the methodologies cannot be used without predicting the pandemic when the NPIs are adjusted. Besides, different sets of NPIs enforced in different places at different times often come with different effects^10, 16^. Without considering NPIs’ varieties on space, timing, and duration, we cannot understand whether these NPIs have had the desired effect of controlling the epidemic. In this study, we aim to tackle the challenge by modeling the COVID-19 spreading as a feedback control system, where the NPIs and vaccination note the controllers, and then to help policymakers determine the magnitude and timing of interventions’ deployment as circumstances change.

Engineering perspectives are useful in epidemic modeling^19^, including the principle of control theory that provides a theoretical basis for NPIs’ functioning and transmission management. Control theory, originally developed for engineered systems with applications to power grids, manufacture, aircraft, satellite, and robots, has recently been adapted to understand the controllability of complex systems emerging in ecology, biology, and society. The recent work about network control enables us to identify the minimal driver nodes^20^ or lowest control costs^19, 21^ for node control, edge control^22^, target control^23^, multilayer control^24, 25^, temporal control^26^, and data-driven control^27^. However, we continue to lack general answers to practically applying control theory to human and natural systems, like the dynamical system for disease spreading. The difficulty is rooted in the fact that we know the pedals and the steering wheel are the drivers prompting a car to move with the desired speed and in the desired direction, but the practical drivers are unknown for complex human and natural systems^28, 29^. Specifically, when focusing on the COVID-19 pandemic, we hypothesize that non-pharmaceutical and pharmaceutical interventions are the “drivers” to determine the dynamics of the COVID-19 pandemic in each location through controlling the infection, recovery, and death rates^29^. We validate this hypothesis by developing a parsimonious model that excellently predicts how the interventions influence the spreads in 381 MSAs in the US and ultimately estimates the end of COVID-19.

## Results

### Model COVID-19 spreading as a feedback control system

Increasing evidence shows that the spread of COVID-19 follows compartmental models^10, 12, 30–32^, such as the SIRD (Susceptible-Infectious-Recovered-Death) model, which is mathematically described by the nonlinear equations expressing a population balance as follows^33^:

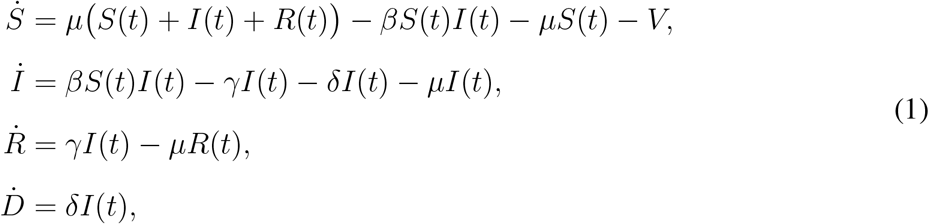

where *S, I, R*, and *D* are the susceptible, infected, recovered and death numbers, respectively. *S* + *I* + *R* + *D* = Ω and Ω is the population at the given place. *V* is the ratio of people full vaccinated with efficacy rate 90%^34^. The parameter *µ* is the crude birth and death rate, and epidemiological parameters *β, γ*, and *δ* are the infection, recovery and death rates (Θ = [*β, γ, δ*]^*T*^). Studies show that epidemiological parameter are time-dependent, which adapt accordingly to the change of interventions^12, 13^. By defining *β*_0_, *γ*_0_, and *δ*_0_ as the infection, recovery and death rates before interventions are imposed (Θ_0_ = [*β*_0_, *γ*_0_, *δ*_0_]^*T*^), we define Θ(*t*) = Θ_0_ + *U*_Θ_(*t*), where *U*_Θ_(*t*) = [*U*_*β*_(*t*), *U*_*γ*_(*t*), *U*_*δ*_(*t*)]^*T*^ is a vector of the control input signals. As shown in Fig. 1a, the controllers *U*_Θ_(*t*) work as the edge control^22^ and the vaccination *V* works as the node control^20^. Based on nonlinear feedback control law, we develop the controllers *U*_Θ_(*t*) using the feedback linearization approach. Then the SIRD model could perfectly track the reference trajectories which is generated from reported real-world pandemic data. As illustrated in Fig. 1b, the output model trajectory fits the real-world 3-dimensional data when the time-varying controllers are included, while it fails to fit when the controllers are not considered. Note that our approach is general and can be extended to other models that consider *n*-dimensional data.

**Figure 1:**
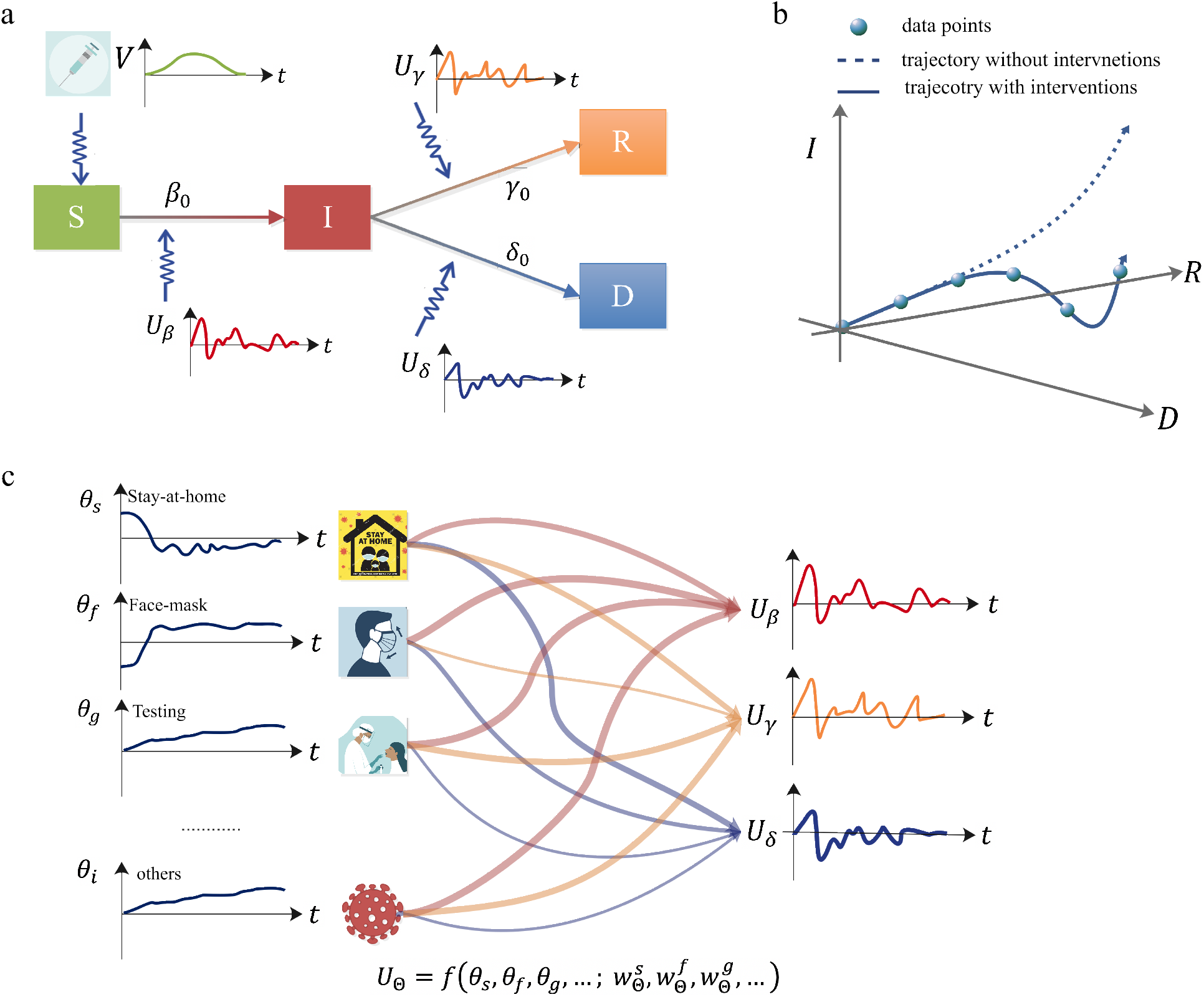
The SIRD-model based feedback control system reveals how the interventions’ magnitudes and timing govern the dynamics of disease. **a** Based on nonlinear feedback control law, we develop the controllers *U*_Θ_ = [*U*_*β*_, *U*_*γ*_, *U*_*δ*_]^*T*^ working as the edge control on infection rate, recovery rate, and death rate. On the other side, the vaccination *V* works as the node control on susceptible population. **b** With these controllers, the output trajectory of disease *X* = [*S, I, R, D*]^*T*^ of the feedback control system fit withe real-world infection data of the disease. **c** Through the model, which includes the nonlinearity or interactions between NPIs, the human behaviour toward NPIs (e.g., stay-at-home order *θ*_*s*_, face-mask wearing *θ*_*f*_, and testing *θ*_*g*_) are linked to the controllers *U*_Θ_ through their effects (e.g., 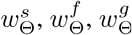).

Next, we propose the parsimonious model by employing the *difference-in-difference* estimation, which maps human behavior toward NPIs into the designed controller, as shown in Fig. 1c. This method enables us to measure the effect of NPIs on the controllers *U*_Θ_ by comparing the infection dynamics before and after the same region’s NPIs deployment. Beyond existing models, which assume the effects on policies are approximately linear^10^, we also identify the interactions between policies. Thus, we are able to compile the evolution of human behaviour toward the NPIs, ***θ***_*I*_ = [*θ*_*s*_, *θ*_*f*_, *θ*_*g*_]^*T*^, (e.g., stay-at-home order *θ*_*s*_, face-mask wearing *θ*_*f*_, and testing *θ*_*g*_) to the evolution of the control signals with their respective effects, 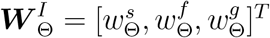.

In short, our two-steps approach captures how the NPIs and vaccinations govern the disease dynamics, through combining the principles of control theory^29, 35^ and statistical estimation^10, 36^.

### Tracking the pandemic’s trajectory with designed controllers

To validate its accuracy in predicting the disease evolution in the future and its applicability in determining the needed interventions to control the infection process, we test the NPIs data and infection data at 381 Metropolitan statistical areas (MSAs) from 1 April 2020 to 20 February 2021. The MSAs, defined as the core areas integrating social and economic adjacent counties, is contiguous areas of relatively high population and traffic density. We consider each MSA as a “closed population” the disease evolution could be modeled by the nonlinear SIRD dynamical model, Eq. (1), in a compact form, as

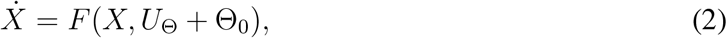

with *X* = [*S, I, R, D*]^*T*^ is the state/output vector and *U*_Θ_ = [*U*_*β*_, *U*_*γ*_, *U*_*δ*_]^*T*^ is the input vector. Using feedback linearization control design, we construct a nonlinear feedback control law 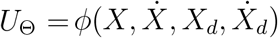, to track the real-world pandemic trajectories *X*_*d*_ = [*S*_*d*_, *I*_*d*_, *R*_*d*_, *D*_*d*_]^*T*^ (see Eqs. (6-11) in Methods). The feedback law *ϕ*(.) relies on the measurements of the model full state *X* and the reference trajectory (*X*_*d*_) and their time derivative (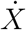 and 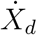).

Theoretically, for each MSA, the output trajectory *X* perfectly fits the reported infection trajectory *X*_*d*_ when the control action *U*_Θ_ is applied. This fact is illustrated in Fig. 2, which shows the evolution of the predicted and reported trajectories from 1 April 2020 to 20 February 2021 for two MSAs, namely the “New York” MSA (with New York as the core) and ‘Houston’ MSA (with Houston as the core). Moreover, the excellent alignments between the real-world and model data of all 381 MSAs indicate our approach’s predictive power, as shown in Fig. 2b. On the other side, Fig. 2c shows all MSAs’ infection rate [*β*_0_ + *U*_*β*_(*t*)], recovery rate [*γ*_0_ + *U*_*γ*_(*t*)], and death rate [*δ* + *U*_*δ*_(*t*)], respectively, and mark out the respective rates for the two examples of MSA. Overall speaking, the infection rate and recovery rate decrease till May, rebound in June, decrease again to the lowest in September and start to fluctuate till February. The death rate continues to decrease in October and stays relatively constant beyond. As validated in literature^4^, “New York” MSA enforces interventions more effectively and earlier, leading to a relatively lower infection rate, recovery rate, and death rate than most MSAs till October. One can observe that “Houston” MSA follows medium patterns.

**Figure 2:**
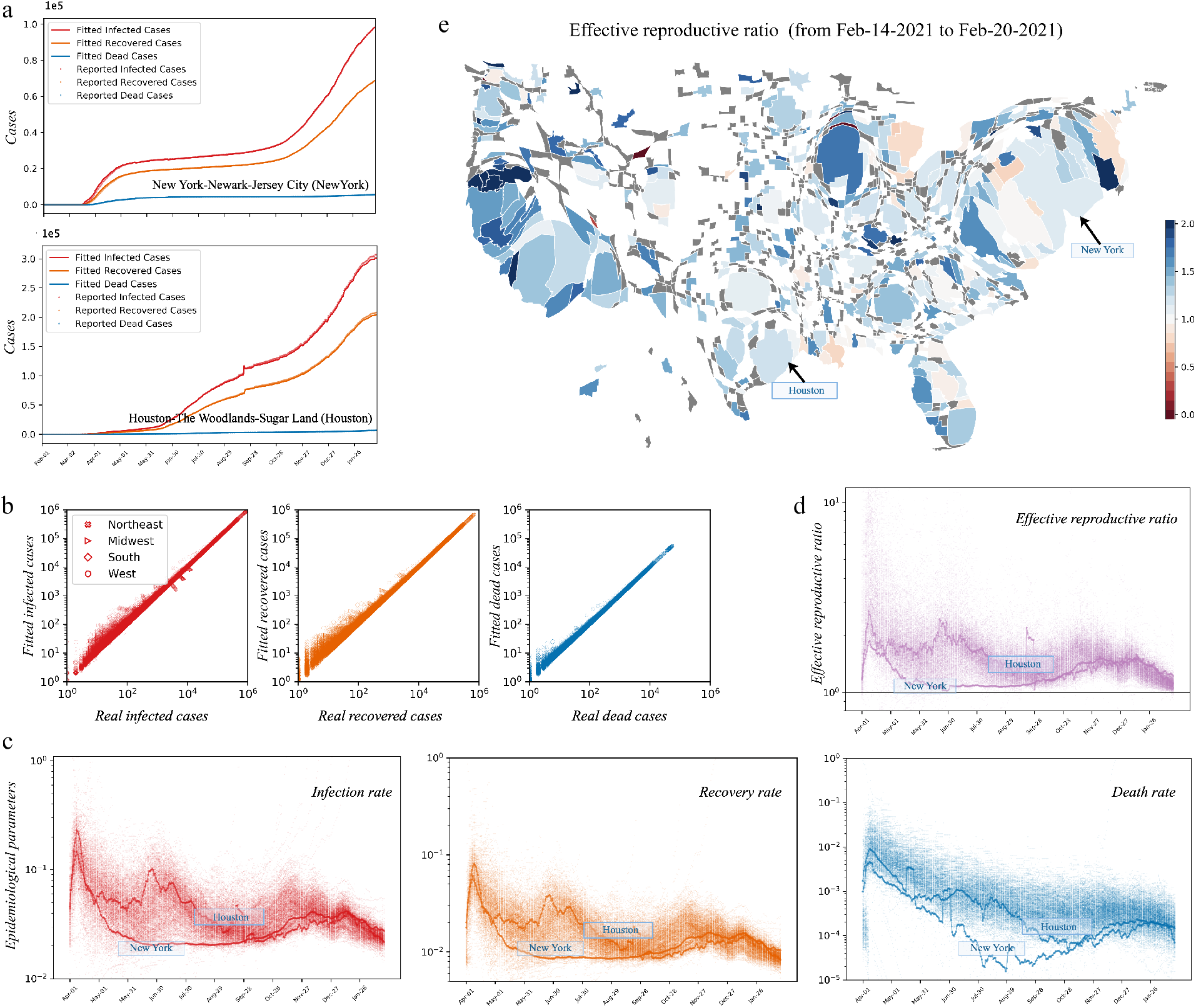
The designed feedback controllers *U*_Θ_ drive the output trajectories 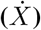 in the SIRD model ingeniously fits the reported trajectories 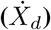 for 381 MSAs. **a** The reported data (dots) of infected/recovered/dead cases are fitted with the output trajectory (solid line) for example MSAs, i.e., “New York” MSA and “Houston” MSA. **b** Comparison between the reported data and output data at all MSAs. Each MSA represent a dot. **c** All MSAs’ temporal infection rate, [*β*_0_ + *U*_*β*_(*t*)], recovery rate, [*γ*_0_ + *U*_*γ*_(*t*)], and death rate, [*δ*_0_ + *U*_*δ*_(*t*)] with marking out the examples MSAs’ rates. **d** shows the MSA’s temporal effective reproductive ratio (*R*_*e*_) and **e** shows each MSA’s average effective reproductive ratio from 14 February 2020 to 20 February 2021 in the cartogram map, in which geometry of regions are distorted according to their population. The effective reproductive ratio reaches the lowest as of 20 February 2021.

Having the daily transmission rate, recovery rate, and death rate, we compute the effective reproductive ratio 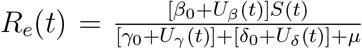;, representing the expected number of secondary infected cases at time *t* when no vaccinations are rolled out. The function *R*_*e*_(*t*) is a critical threshold in understanding whether the outbreak is under control. More precisely, if *R*_*e*_(*t*) *<* 1, then the ongoing outbreak will eventually fade out, whereas *R*_*e*_(*t*) > 1 means an acceleration of the infection dynamics leading to a substantial growth of infected cases and deaths. Fig. 2d-e respectively visualize the temporal and spatial distribution of the MSAs’ effective reproductive ratio. Our results reveal that just a few MSAs’ effective reproductive ratios ever reach *R*_*e*_(*t*) *<* 1, implying the need to implement more rigorous interventions to achieve an effective controlling of the pandemic. For example, from 14 February 2021 to 20 February 2021, the average effective reproductive ratios in “New York” and “Houston” MSAs are evaluated as 1.468 and 1.393, respectively, revealing a critical need for stronger interventions.

### Mapping human behaviours as actuators to steer NPIs as controllers

Although our controller, *U*_Θ_, precisely predicts the infectious, death toll, and recovery, it is thus far unknown how the measurable interventions change the controllers. Compared with the driving car, it is similar that we know the desired speed and direction in order to move the car to the target location, but we do not know what the pedals and the steering wheel for epidemic control are and how the changes in the pedals and the steering wheel determine the speed and direction. In another word, as depicted in Fig. 1, we assume that the changes of interventions, directly steering the control signals, will shape the disease dynamics when they are tightened and loosen to different levels. For multiple NPIs ***θ***_***I***_, we divide them into two sets, i.e., the set of community NPIs *ϑ*^*c*^ (e.g., social distancing and quarantine) and the set personal NPIs *ϑ*^*p*^ (e.g., face covering, test, and frequent hand wash). Then we develop the following mathematical model 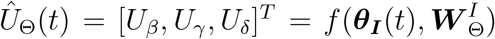 as (see [Eqs. (12-15)] in Methods)

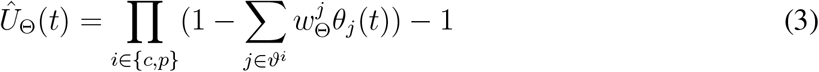

where *Û*_Θ_(*t*) is the estimate of the control action based on NPIs with their magnitudes *θ*_*j*_(*t*) and their impact value 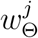. For a the specific NPI *j*, large 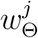 value demonstrates that NPI *j* has strong impact. If 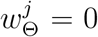, the control *Û*_Θ_(*t*) is independent with the NPI *j*. The term Π _*i*∈{*c,p*}_ indicates that community NPIs *ϑ*^*c*^ and personal NPIs *ϑ*^*p*^ have a joint affect on the controller. For either community NPIs *ϑ*^*c*^ or personal NPIs 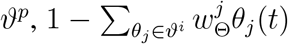 term denotes the combined impact of the NPI set. Usually, *Û*_Θ_ is a non-positive value, showing the reductions in each controller.

Given the collected data sets for eight NPIs, we evaluate the goodness of fit versus different combinations of NPIs to find an effective parsimonious model for Eq. 3. A parsimonious model, which has great explanatory predictive power, accomplishes a better prediction of controllers as few NPIs as possible. With 70% of the dataset of NPIs as the training data, we use the mean absolute error to test the predictive accuracy for the rest 30% of the dataset. Fig. 3 shows that including three NPIs in the model gains the highest predictive accuracy for the designed controllers (see supplementary text for the details). We find that the most representative NPIs for Eq. 3 are: (1) stay-at-home order, represented by the normalized ratio of excessive time of staying at home, *θ*_*s*_; (2) face-mask wearing, represented by the fraction of people wearing face masks, *θ*_*f*_; (3) testing, represented by the normalized fraction of tested population, *θ*_*g*_. Commonly, stay-at-home order and face-mask wearing have a positive impact on decreasing the number of reported cases. Differently, testing *θ*_*g*_(*t*) may positively or negatively impact the reported infected cases. The reason is that more testing could allow identifying more cases when the number of testing is not sufficient, and yet, more testing may also lead to less infected cases^6^. Representing the three NPIs as ***θ***_*I*_ = [*θ*_*s*_, *θ*_*f*_, *θ*_*g*_], then,

**Figure 3:**
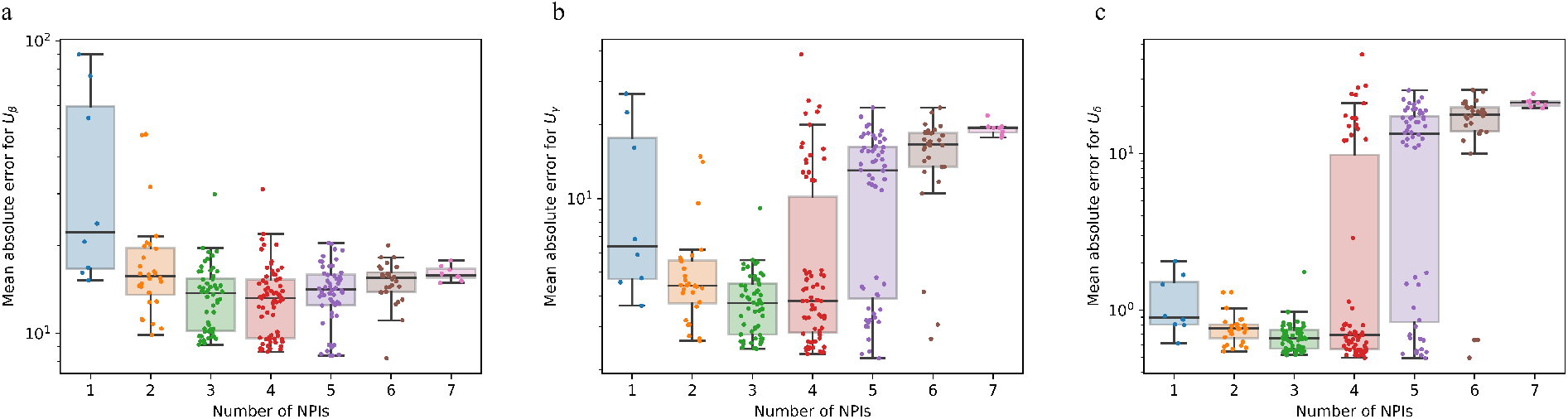
NPIs selection for predicting the designed controllers *U*_Θ_. Given eight NPIs, i.e., stay-at-home order, school closure, quarantine, working from home, face-mask wearing, testing, frequent hand wash, and avoid crowding, we test the accuracy of predictive model of Eq. (4) with different combinations of NPIs for *U*_*β*_ (**a**), *U*_*γ*_ (**b**), and *U*_*δ*_ (**c**). The model achieves high parsimony (with fewer NPIs) and high level of goodness of fit (with lowest mean absolute error) using three NPIs, which are stay-at-home order, face-mask wearing, and testing.

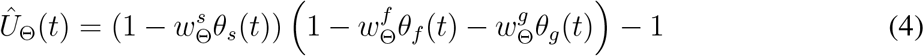

In the following studies, we only use these three selected NPIs for predictions.

### Effects of NPIs on shaping the disease dynamics

Based of the parsimonious model of Eq. (4), we learn the parameter-by-intervention specific marginal effects 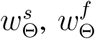, and 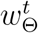, reflecting the variations of *Û*_Θ_ as a function of the evolution of the time-dependent local NPIs *θ*_*s*_(*t*), *θ*_*f*_ (*t*), and *θ*_*t*_(*t*) at MSAs. With the objective to directly infer the *Û*_Θ_ with the NPIs data, we first use 70% dataset of NPIs as the training data to learn the marginal effects. Then, we estimate the counterparts *Û*_Θ_ given as Eq. (4) using the 30% left testing datasets and evaluate the fit between *U*_Θ_ and *Û*_Θ_. Taking the “New York” MSA and “Houston” MSA shown in Fig. 4a-b as illustrative examples; given the evolution of NPIs, the estimated control signals *Û*_*β*_ and *Û*_*δ*_ with the learned marginal effects, fit the predicted model-based control law defined in terms of *U*_*β*_ and *U*_*δ*_. Based on the proportionality between *U*_*γ*_(*t*) and *U*_*β*_(*t*) (see Fig. S2), i.e., *U*_*β*_(*t*)*/U*_*γ*_(*t*) = 2.664, here, we only consider *U*_*β*_(*t*) and *U*_*δ*_(*t*).

**Figure 4:**
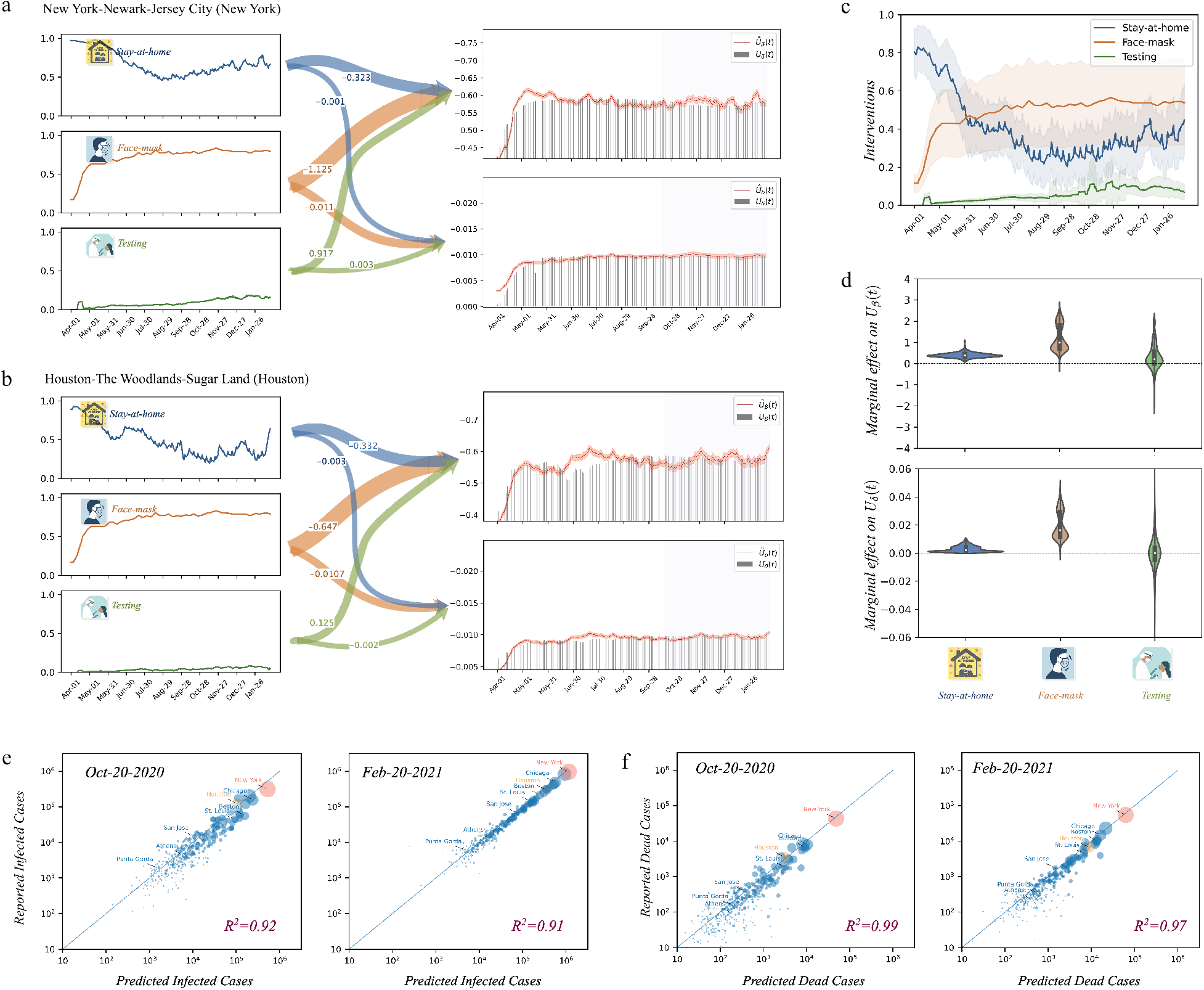
The learned effects enable to use magnitude of NPIs to predict infection at each MSA. Using the 70% NPIs and infection data, we learned the NPIs’ effects in the model of Eq. (4). Based on the learned effects, **a**,**b** illustrates of how the magnitude of NPIs determine the controllers (*Û*_*β*_ (*t*) and *Û*_*δ*_ (*t*)) in the example MSAs. To test the accuracy of model, we compare [*Û*_*β*_ (*t*) and *Û*_*δ*_ (*t*)] with analytical controllers [*U*_*β*_(*t*) and *U*_*δ*_(*t*)] with the left 30% NPIs and infection data (vertical shaded area) in the right-side plots of **a**,**b. c** All MSAs’ magnitudes of NPIs. The solid line represents the average, and the shaded area represents the standard variance for all MSAs. Though large difference between MSAs’ magnitudes of interventions, in **d**, the marginal effects for stay-at-home order and face-mask wearing are mainly negative. Testing, which may helps to find more infection and death, has either negative or positive effect. Thus, by taking the estimated controllers *Û*= [*Û*_*β*_, *Û*_*β*_, *Û*_*γ*_] as the input the SIRD model, we find the estimated infection/death assemble with reported infection/death with *R*^2^ > 0.9, see the two example dates in **e** and **f**. Fig. S3 shows the estimation accuracy for all others dates. It should be noted that as the *Û*_*γ*_ keeps constantly proportional to *Û*_*β*_ (see Fig. S1), there is no need of further exploring *Ûγ*.

It is remarkable that for all MSAs, Eq. (4) stays robust to the huge heterogeneity of locality. As depicted in Fig. 4c, the NPIs have a great high standard deviation (*σ*) across MSAs, especially for stay-at-home order and face-mask wearing. One can notice that the average magnitude of stay-at-home order decreases from 80% to 40% recently with *σ* = 0.140. Besides, nearly 53.56% of people are wearing face masks when being outdoors with *σ* = 0.20 while the average magnitude of testing increases to 9.19% with *σ* = 0.032. To test the robustness of the model of Eq. (4), we trained and validated the model iteratively on different MSAs’ datasets for NPIs. The distributions of marginal effects across the three NPIs are shown in Fig. 4d. Most MSAs’ marginal effects for stay-at-home order and face-mask wearing are negative. The statistics imply the generic feature of the Eq. (4) in capturing NPIs’ effects despite the huge regional heterogeneity of human behaviours. However, the average marginal effect of testing has two opposite outcomes. For *Û*_*β*_ (*t*), 73.5% of MSAs’ testing’ marginal effects are positive, meaning 26.5% of them are negative. For *Û*_*δ*_ (*t*), 46.0% of MSAs’ testing’ marginal effects are positive, meaning 54.0% of them are negative. The positive effects suggest that testing is favorable to finding more infections and deaths, while the negative effects show that testing reduces infection or death.

Applying the estimated control signals *Û*_*β*_ (*t*), *Û*_*δ*_ (*t*), and *Û*_*γ*_ (*t*) = *Û*_*β*_ (*t*)*/*2.264 to the SIRD model, we find that the new output trajectories 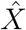 fit the reported infection *X*_*d*_ with *R*^2^ ≥ 0.9, as show in Fig. S3. As well, Fig. 4e-f depict the predicted infected/dead cases with reported infected/dead cases on 20 October 2020 and 20 February 2021. All the results further validate the model in assessing NPIs’ effects on disease dynamics.

### Needed NPIs and vaccinations for achieving *R*_*e*_ to manageable level

The two-steps approach, integrating the designed controllers in Eqs. (1-2) and the effect of NPIs on controllers in Eqs. 4, has successfully mapped human behaviors to NPIs to infection rate, recovery rate, and death rate of the SIRD model. Then, the effective reproductive number *R*_*e*_ can be equivalently translated in terms of magnitudes of NPIs (***θ***_*I*_) and *V* (the ratio of people full vaccinated with efficacy rate 90% ^34^), that is,

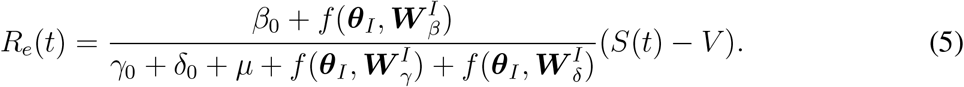

From the equation above, we can determine the needed magnitude of NPIs for achieving *R*_*e*_(*t*) *<* 1, criteria for the disease die out, under a given vaccination coverage. Here, the vaccination coverage is viewed as an open-loop control action or a given parameter whose assigned value affects the speed of propagation of the disease.

When there is no vaccination administered in the US (like before 12 January 2021 *V* = 0), the needed magnitude of NPIs to achieve an effective reproductive number below the threshold, *R*_*e*_(*t*) *<* 1^37^, for ‘New York’ and ‘Houston’ MSAs are shown in Fig. 5a-b. The “triangular prism” in three dimensions represents the needed magnitude of the stay-at-home order, face-mask wearing, and testing. The horizontal slices of the “triangular prism” are the visualization for the needed magnitude of stay-at-home order and face-mask wearing if the magnitude of testing is fixed. If stay-at-home order and face-mask wearing interventions are enforced at a level greater than about 80%, *R*_*e*_(*t*) *<* 1, which forms a “triangle” as shown in Fig. 5a-b. As opposed to stay-at-home order and face-mask wearing, the testing intervention has a positive marginal effect on ‘New York’ MSA and ‘Houston’ MSA. Hence, in both cases, the “triangle” becomes smaller for larger testing. Some MSAs (‘Log Angles’ MSA and ‘Miami’ MSA in Fig. S4) have a negative marginal effect for testing intervention, and their “triangle” becomes bigger for larger testing. According to their testing interventions ‘ marginal effects, the shapes of “triangular prism” for more MSAs are shown in Fig. S4.

**Figure 5:**
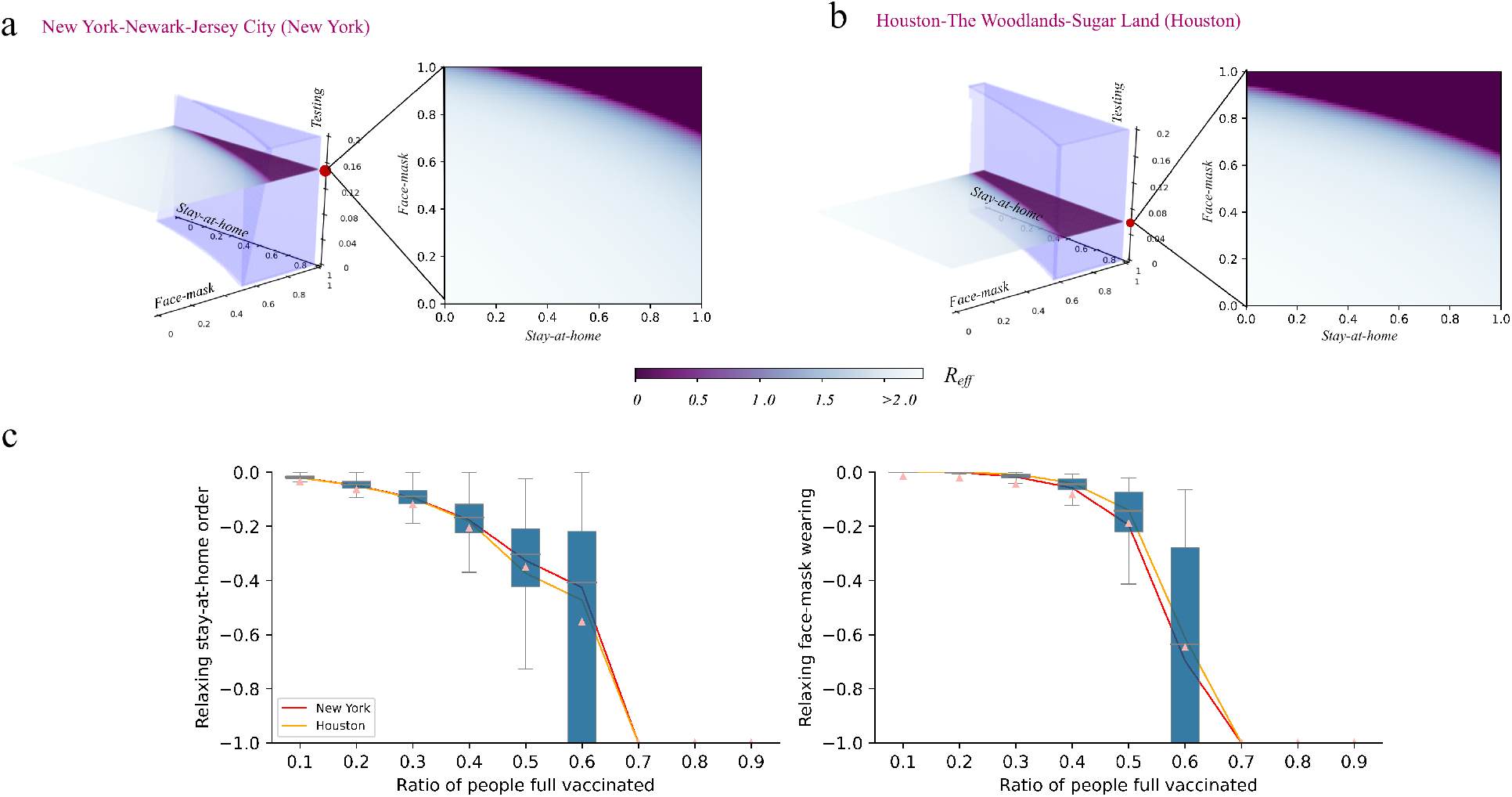
Needed NPIs to keep effective reproductive ratio *R*_*e*_ ≤ 1 with difference vaccine coverage. Take the “New York” MSA and “Houston” MSA as the examples; **a**,**b** show the needed magnitude for NPIs in order to keep *R*_*e*_ ≤ 1 with zero vaccine coverage. The horizontal slices of the “triangular prism” are the visualization for the needed magnitude of stay-at-home order and face-mask wearing if the magnitude of testing is fixed. **c** When people are full vaccinated at different level (vaccine coverage), how much extents of stay-at-home order and face-mask wearing could be relaxed while keep *R*_*e*_ = 1. The magnitude of testing is fixed as each MSA’ recent testing capacity. These results demonstrate relaxing interventions need thorough and careful considerations when the ratios of full vaccinated people *<* 0.6.

Since 12 January 2021, the US began the first jab of vaccine, and undoubtedly, *R*_*e*_(*t*) could be smaller with mass vaccinations according to the Eq. (5). For different ratios of fully vaccinated people with 90% efficiency, we could find at least how much NPIs could be relaxed (reduced) to achieve *R*_*e*_(*t*) = 1 in Fig. 5c. Setting aside testing intervention, more magnitude of stay-at-home order and face-mask wearing interventions could be eased if more people are fully vaccinated. However, when 60% of people are fully vaccinated, only 55.3% of stay-at-home order and 64.8% face-mask wearing could be eased. When 70% people are fully vaccinated, the stay-at-home order and face-mask wearing could be eased entirely. However, to achieve *R*_*e*_(*t*) *<* 1, the policymakers should relax NPIs with more cautions.

### Needed days of eliminating the pandemic locally with vaccinations

Moreover, due to the unknowns around mutations in escalating the disease’s infectiousness^38^ and human’s long-term immunity to the disease^39^, suppressing the disease to an acceptable level with NPIs but not aiming to eliminate it could make COVID-19 an endemic^40, 41^. A COVID endemic could claim millions of more lives each year and cause devastating economic burdens on immunization, treatment, and prevention. Also, repeated tightening and loosening interventions due to recurrent outbreaks, without a doubt, increase the difficulty to decide the right moment of enforcing exit strategies^14, 35, 42, 43^. Currently, three vaccines are authorized and recommended in the United States from Pfizer-BioNTech, Moderna, and Johnson & Johnson. By 10 March, more than 37.4 million adults in the U.S. have been fully vaccinated^44^. Despite continued efforts of NPIs and vaccine roll-out, the plateaued numbers of infection cases and the fast-spreading variants still pose significant uncertainty about whether “zero COVID” could be achieved locally in the US. Given the examples of New Zealand^45^, Vietnam, Brunei, and Island states in the Carbbean^40^, which ever reached a “Zero COVID” stage, we investigate the possibility of reaching zero contamination in the MSAs. We refer to “Zero COVID” to describe a situation that leads to no new cases at least for three months in a given location, referring to literature^46^. Furthermore, the day when the following three months having zero new cases is defined as the elimination day.

We assume that the ratio of fully vaccinated people (also call vaccination coverage), follows the innovation adoption model according to Rogers^47^, which is a normal distribution, *H*(*t*), see Eq. (16)-(17). Then, the total number of immunized people is defined as *V* (*t*) = *aH*(*t*) where *a* is the vaccine effectiveness (*a* = 90%). As shown in Fig. 6a, the reported data reveal that the ratio of fully vaccinated people from 12 January 2021 to 20 February 2021 in the US increases to 5.40%. By fitting the real-world ratio, we obtained the vaccination coverage as a curve that gradually reaches saturation within different periods (*T*_*s*_). Considering a full vaccination of 50% of people after 360 days since 12 January 2021, the average elimination day for all MSAs corresponds to the hundred seventieth day. Under this configuration, from in Fig. 6b, one can notice that ‘New York’ MSA reaches a total elimination stage after 197 days while ‘Houston’ MSA achieves a total elimination stage after 187 days. Of course, one could define a vaccination strategy rollout accounting for nonuniform saturation coverage in different periods. Fig. 6c comprehensively presents all MSAs’ elimination days for vaccination coverage ranging from 0% to 90% with vaccination period (*T*_*s*_) equals to 90 days, 180 days, 270 days, and 360 days. Our results reveal that broader vaccination coverage further reduces both the remoteness of the elimination day and the death toll. On the other side, a larger vaccination period expands the time range needed to reach the elimination day and leads to a more significant death toll. For the case in Fig. 6b, a total of 23,920,837 more people would be infected, and 1,199,202 more people would be dead because of COVID-19 if 50% of people are fully vaccinated in 360 days. On the contrary, if 90% of the population is fully vaccinated in 360 days, the average elimination day for all MSAs is about 129 days after implementing the vaccination strategy, with 20,428,296 more infected cases and 9,127,85 more dead cases overall. As shown in Fig. S6-S7, “New York” MSA, ‘Los angels’ MSA, “Chicago” MSA, “Dallas” MSA, “Phoenix” MSA, “Boston” MSA and “Philadelphia” MSA have far more death than other MSAs, meaning they need stricter interventions in reducing death because of COVID-19. In summary, all these results emphasize that the earlier, aggressive vaccination strategies are deployed in each MSA, the earlier the pandemic could be eliminated, leaving fewer deaths from the disease.

**Figure 6:**
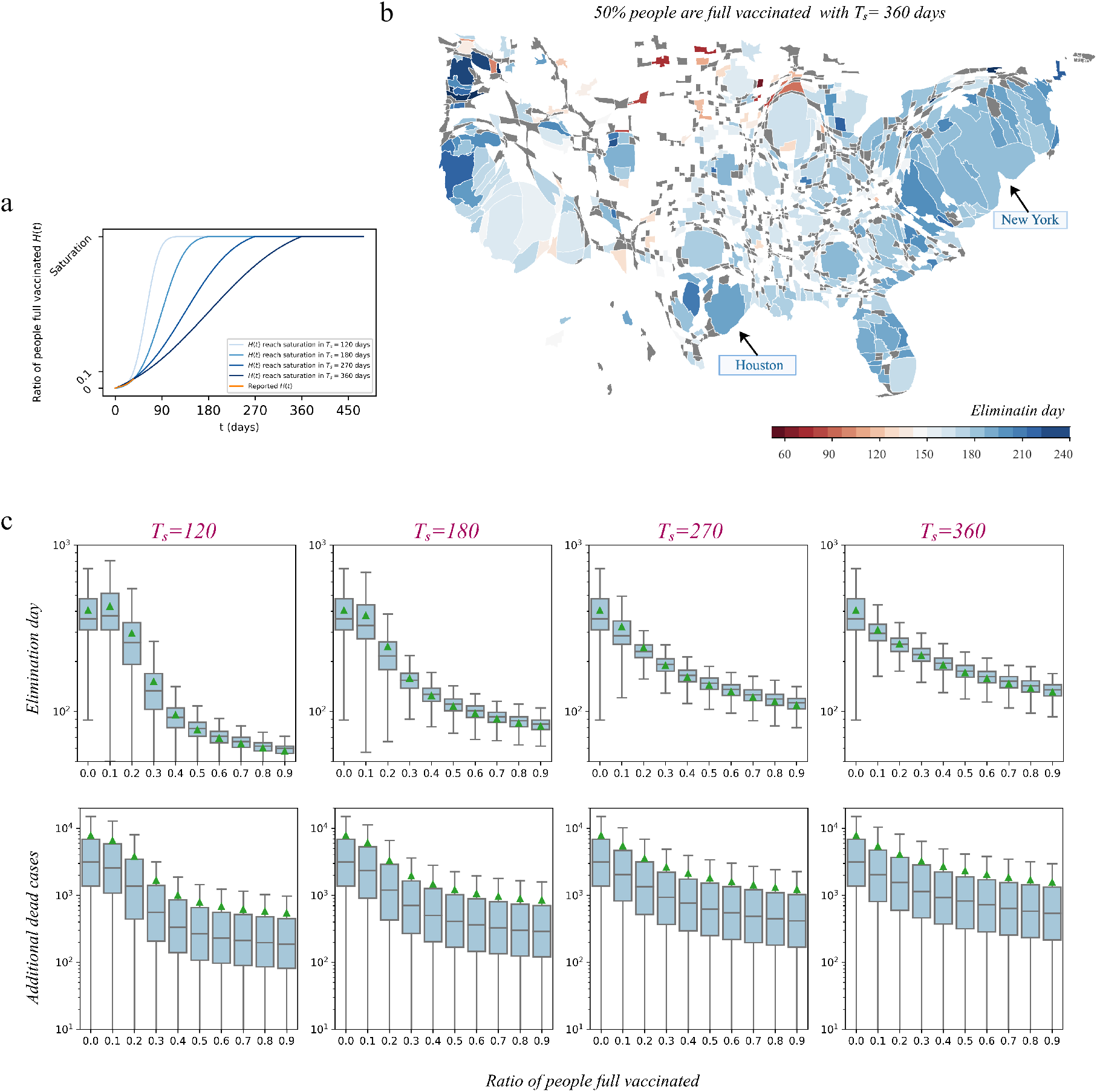
Days needed for reaching “Zero COVID” if the saturation of vaccination coverage are reached within 90 days, 180 days, 270 days, and 360 days. By (1) defining “Zero COVID” as that the disease is eliminated when there are zero new infection for 90 days, (2) defining the day reaching “Zero COVID” as the elimination day, (3) assuming that interventions (i.e., stay-at-home order, face-mask wearing, and testing) keep same as that from 5 January 2021 to 11 February 2021, (4) consider ratio of people full vaccinated as the vaccination coverage, we test the elimination days for MSAs since 12 January 2021. **a** The vaccination adoption model *H*(*t*) illustrate how saturation of people full vaccinated is reached in period *T*_*s*_. Specifically, the orange line is the reported cumulative vaccination coverage. **b** MSAs’ estimated elimination days if 50% are full vaccinated with 360 days. **c** Box plots for the elimination days, and the additional death till elimination days for different saturation of vaccination coverage within different time period *T*_*s*_.

## Discussion

We present the COVID-19 pandemic as the feedback control system to show how the evolution of disease adapt to human behaviours to NPIs and the vaccine coverage. This approach enable us to model the linear and nonlinear effects of multiple NPIs on the SIRD model-based feedback controllers on the epidemiological parameters. Through reducing eight NPIs to three representative three (i.e., stay-at-home order, face-mask wearing, and testing), we obtain the model having great explanatory predictive power toward disease dynamics. By studying the NPIs at 381 MSAs from 1 April 2020 to 20 February 2021, we find that the two-steps approach is robust and efficient to predict daily infection and death accurately. Beyond in line with existing studies that NPIs are effective in suppressing the disease outbreaks^30, 48, 49^, we could directly link the NPIs’ marginal effects to the effective reproductive number *R*_*e*_. It implies that, without the need for up-to-date knowledge of current infections and ‘nowcasting’. Besides accurate forecast on both case counts and deaths, this approach could provide practical information for policymakers regarding the extent of safely relaxing NPIs and the needed vaccination coverage to end COVID-19 locally. The approach is also universal and thus can be used by policymakers elsewhere.

Through analyzing the needed NPIs for keeping *R*_*e*_ *<* 1, we find that MSAs should continue to enforce their stay-at-home order and face-mask wearing. Loosening the degree below 80% could lead to a resurgence of COVID-19. Our results show that MSAs should continue to require wearing masks and unless 60% of people are fully vaccinated, the MSA should not relax the stay-at-home order and face-mask wearing intervention.

There is an ongoing debate on if the elimination of COVID-19 (”Zero COVID”) is possible^40, 41^. It is considered by some as the only way to prevent future crises and cannot be achieved without mass vaccination. Using the “Zero COVID” as a guiding criterion, i.e., no new infection at least for three months, we test how many days it can be achieved if 50% MSA populations are vaccinated in a 360-day period. We find that, on average, it takes 170 days to reach the “Zero COVID” milestone. It is worth noting that the promising result can be undermined by many external factors, like importation cases from other areas, making the total elimination a long process with fluctuations on new infections. In addition, some scientists argue pursuing the “Zero COVID” comes at a huge cost to normal human life^40, 41^ and can thus out-weigh the benefits. Nevertheless, our experiments reinforce the argument that maintaining NPIs and encouraging people to get vaccines are necessary and key strategies to lower the new infections as much as possible.

Our study has some limitations. The first limitation is rooted in the dataset for COVID-19 and the dataset for NPIs. Given the mass mild or asymptomatic infections, the inaccuracy of reported infections and deaths would increase the uncertainty of the SIRD model-based feedback controllers. The second limitation is assuming each MSA as the closed population, which ignores the fluctuation of infections caused by case importation and exportation. This simplification would influence the results from needed NPIs and PIs for suppressing COVID-19 to elimination test. The third limitation is assuming vaccination adoption follows the curve of diffusion of innovations without considering people’s attitude to vaccinations. As the attitudes towards vaccination vary by age, race, ethnicity, and education, it is hard to capture the full complexity. In our current work, we use feedback linearization to design the control signals, and we may improve the accuracy by considering other controller design strategies, for example, adaptive controller^50, 51^, model predictive control^52^, or intelligent control^53^. Nevertheless, our study provides practical insights into tightening or relaxing NPIs for the aim of living with COVID-19. Also, we provide the possibility of achieving “Zero COVID” in metropolitan areas if vaccination is stable and efficient enough.

## Methods

### Dataset

#### 2019-COVID pandemic

We acquire the second-administrative units’ COVID-19 cases from the 2019 Novel Coronavirus Visual Dashboard operated by the Johns Hopkins University Center for Systems Science and Engineering^54^. It covers the counties’ reported infected and dead cases at the US and the whole countries’ infected and dead cases from 21 January 2020 to 20 February 2021. We integrate counties’ COVID-19 cases for 381 MSAs defined by the United States Office of Management and Budget (OMB), which serves as a high degree of social and economic core areas.

#### Testing Capacity

The testing data at each county is also collected by the Johns Hopkins University Center for Systems Science and Engineering^54^. By integrating the counties’ testing data, we get each MSA’s daily ratio of total testing over MSA’s population.

#### Stay-at-home Data

Daily data about the observed minutes at home and observed minutes outside of home for all devices are counted at each county by the Safegraph^55^, which is a platform of collecting the points of interests (POIs) from anonymous mobile devices. Integrating the data, we could get each MSA’s ratio of time at home, i.e, the fractions of the observed minutes at home over the sum of observed minutes at home and outside home, which is reflective of people mobility pattern for local governments’ anti-contagious policies. By subtracting the benchmark ratio (the average ratio of time at home before 13 March 2020) and then normalizing the ratios from 0 to 1, we get each MSA’s ratio of excessive time at home.

#### People Wearing Face Mask

We assume that the search of “face mask” collected by Google Trend equals to the fraction of people having the awareness of wearing face masks in each MSA. Combined the fractions and the weekly percentages of people wearing face mask in the US provided by YouGov^56^, we could obtain each MSA’s daily percentage of people wearing face mask. We use the similar approach to collect the data of people support school closure, quarantine, working from home, frequent hand wash, and avoid crowding.

#### Vaccination Data

CDC provides the overall US COVID-19 Vaccine Distribution and Administration, mainly for Pfizer-BioNTech and Moderna.

### Deriving the feedback controllers

The feedback controllers measure the output of the SIRD model and then manipulates the inputs on infection rate, recovery rate, and death rate of as needed to drive the model output toward the desired COVID-19 pandemic trajectory. In another word, given the controllablility of the SIRD model (see supplementary text), then we could rewrite the Eq. (1) as

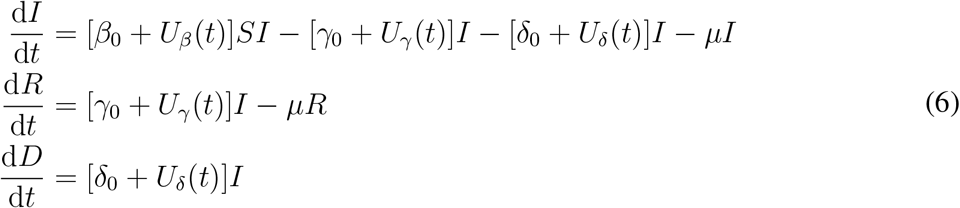

with the controller set *U*_Θ_(*t*) = [*U*_*β*_, *U*_*γ*_, *U*_*δ*_]^*T*^. *U*_*β*_, *U*_*γ*_, *U*_*δ*_ are the controllers on infection rate, recovery rate, and death rate. For the reference COVID-19 pandemic trajectory *X*_*d*_ = [*S*_*d*_, *I*_*d*_, *R*_*d*_, *D*_*d*_]^*T*^ and its corresponding output trajectory *X*, the error dynamics *X*_*d*_ −*X* is governed by the following equations

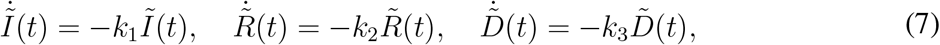

where, 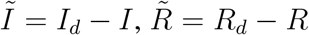, and 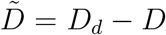. Here, *k*_1_, *k*_2_, and *k*_3_ are positive gains of the designed feedback controller. The solution to the linear ordinary differential equations above are expressed as

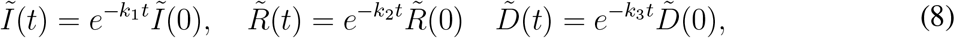

and 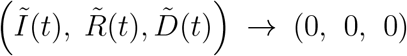. Therefore, the error system is exponentially stable and *X*(*t*) → *X*_*d*_(*t*). The simulation of the closed-loop system is performed selecting *k*_1_ = *k*_2_ = *k*_3_ =0.1. The controllers are

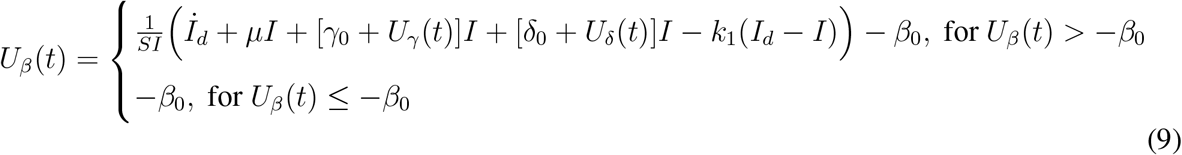

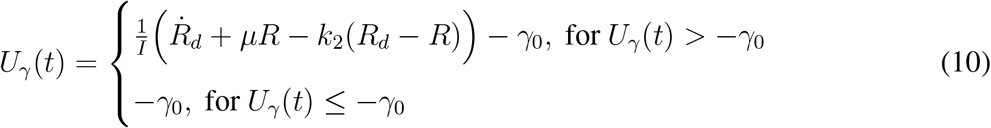

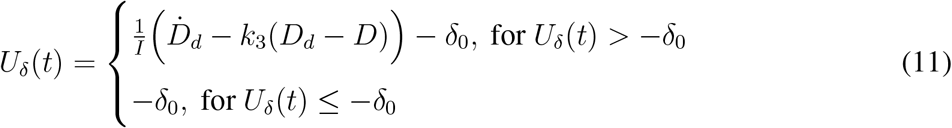

To prevent un-physical negative transmission rate *U*_*β*_(*t*), recovery rate *U*_*γ*_(*t*), and death rate *U*_*δ*_(*t*), the controllers are clipped such that the time-dependent epidemiological parameters are always positive. Here Θ_0_ = [*β*_0_, *γ*_0_, *δ*_0_]^*T*^ is the pre-intervention epidemiological parameters learned by infection of ‘New York’ MSA from 1 March 2020 to 13 March 2020 with Nelder-Mead simplex algorithm^57^. For the controllers, whatever the values for Θ_0_, the error system is stable and *X*(*t*) → *X*_*d*_(*t*).

### Using difference-in-difference method to learn the NPIs’ marginal effects to the controllers

For NPI set ***θ***_*I*_, we use the linear regression model for difference-in-difference to calculate their effects on the vector of infection rate, recovery rate, and death rate *U*_Θ_(*t*) + Θ_0_. Consider the model,

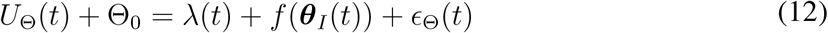

where *λ*(*t*) is the facotr for time trend and *ϵ*_Θ_(*t*) is the residual term. Then, the difference of outcome controllers from time *t* − 1 to time *t* is

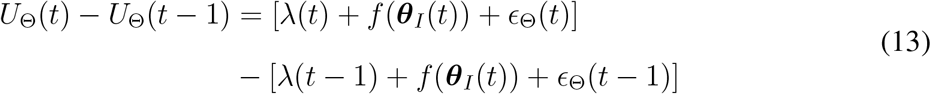

Adding up the all difference from time *t* = 0 to time *t*

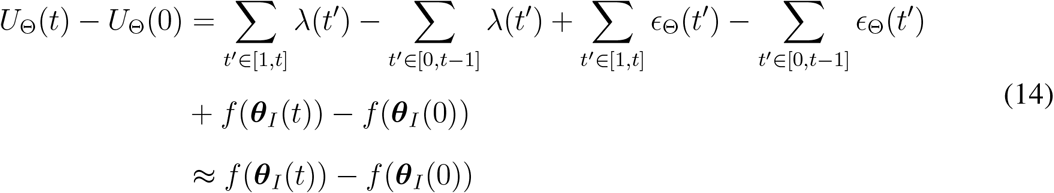

As when *t* = 0 no NPIs are implemented, thus, *U*_Θ_(*t*) = 0 and *f* (***θ***_*I*_(0)) = 0,

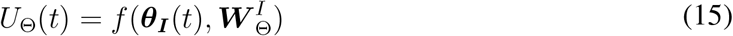

### Vaccination adoption model

Like the innovation adoption model, the daily newly vaccination adoption (vaccination coverage) will follow the bell curve, the normal distribution,

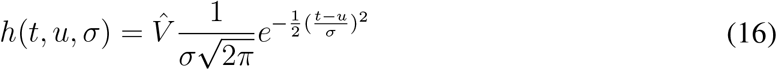

where 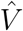is the saturation of vaccination coverage. The the cumulative vaccination adoption (vaccination coverage) is

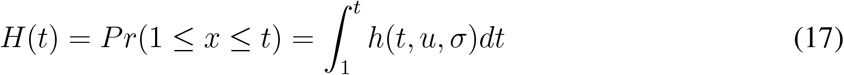

with *t* ∈ [1, *T*_*s*_]. It means, the cumulative vaccination coverage will reach saturation of 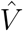 in *T*_*s*_ days. In the test of “Zero COVID”, we tune the *u* and *σ* to fit the real-world vaccination coverage in the United States from 12 January 2021 to 20 Februray 2021 for 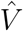 ∈ [0.1, 0.9] and *T*_*s*_ ∈ {120, 180, 250, 360} in Fig. 6.

## Data Availability

Derived Data and codes that support the findings of this article are available in https://github.com/lucinezhong/controllers_ode_COVID.git.

## Acknowledgements

We thank Bhathiya Rathnayake for fruitful discussions on developing the feedback system and the paper.

## Author Contributions

M.D. and J.G. conceived the project and designed the experiments; Q.W. and L.Z collected the data set and analyzed the data; L.Z., M.D., and J.G. carried out theoretical calculations and performed the experiments; all authors wrote the manuscript.

## Data and materials availability

All data needed to evaluate the paper’s conclusions are presented in this paper. Derived Data and codes that support the findings of this article are available in https://github.com/lucinezhong/controllers ode COVID.git.

## Additional information

Supplementary text including Fig. S1-S7, Tab. S1, and Eqs. (S1)-(S12).

## References

1. Organization, W. H. Covid-19 weekly epidemiological update (2020).

2. Interim economic projections for 2020 and 2021. https://www.cbo.gov/publication/56351. Accessed: 2021-02-20.

3. The Council of State Governments. Covid-19: Fiscal impact to states and strategies for recovery (2020).

4. Pei, S., Kandula, S. & Shaman, J. Differential effects of intervention timing on covid-19 spread in the united states. Science advances 6, eabd6370 (2020).

5. Haushofer, J. & Metcalf, C. J. E. Which interventions work best in a pandemic? Science 368, 1063–1065 (2020).

6. Chiu, W. A., Fischer, R. & Ndeffo-Mbah, M. L. State-level needs for social distancing and contact tracing to contain covid-19 in the united states. Nature Human Behaviour 4, 1080– 1090 (2020).

7. Le, T. T. et al. The covid-19 vaccine development landscape. Nat Rev Drug Discov 19, 305–306 (2020).

8. Bubar, K. M. et al. Model-informed covid-19 vaccine prioritization strategies by age and serostatus. Science (2021).

9. Rabby, M. I. I. Current drugs with potential for treatment of covid-19: a literature review. Journal of pharmacy & pharmaceutical sciences 23, 58–64 (2020).

10. Hsiang, S. et al. The effect of large-scale anti-contagion policies on the covid-19 pandemic. Nature 584, 262–267 (2020).

11. Cho, S.-W. Quantifying the impact of nonpharmaceutical interventions during the covid-19 outbreak: The case of sweden. The Econometrics Journal 23, 323–344 (2020).

12. Dehning, J. et al. Inferring change points in the spread of covid-19 reveals the effectiveness of interventions. Science (2020).

13. Chen, Y.-C., Lu, P.-E., Chang, C.-S. & Liu, T.-H. A time-dependent sir model for covid-19 with undetectable infected persons. IEEE Transactions on Network Science and Engineering (2020).

14. Ruktanonchai, N. W. et al. Assessing the impact of coordinated covid-19 exit strategies across europe. Science 369, 1465–1470 (2020).

15. Vardavas, R. et al. The health and economic impacts of nonpharmaceutical interventions to address covid-19. RAND Report TLA 173 (2020).

16. Brauner, J. M. et al. Inferring the effectiveness of government interventions against covid-19. Science 371 (2021).

17. Jing, Q.-L. et al. Household secondary attack rate of covid-19 and associated determinants in guangzhou, china: a retrospective cohort study. The Lancet Infectious Diseases 20, 1141–1150 (2020).

18. Zhong, L., Diagne, M., Wang, W. & Gao, J. Country distancing increase reveals the effectiveness of travel restrictions in stopping covid-19 transmission (2020).

19. Yan, G. et al. Spectrum of controlling and observing complex networks. Nature Physics 11, 779–786 (2015).

20. Liu, Y.-Y., Slotine, J.-J. & Barabási, A.-L. Controllability of complex networks. nature 473, 167–173 (2011).

21. Yan, G., Ren, J., Lai, Y.-C., Lai, C.-H. & Li, B. Controlling complex networks: How much energy is needed? Physical review letters 108, 218703 (2012).

22. Nepusz, T. & Vicsek, T. Controlling edge dynamics in complex networks. Nature Physics 8, 568–573 (2012).

23. Gao, J., Liu, Y.-Y., D’souza, R. M. & Barabási, A.-L. Target control of complex networks. Nature communications 5, 1–8 (2014).

24. Pósfai, M., Gao, J., Cornelius, S. P., Barabási, A.-L. & D’Souza, R. M. Controllability of multiplex, multi-time-scale networks. Physical Review E 94, 032316 (2016).

25. Menichetti, G., Dall’Asta, L. & Bianconi, G. Control of multilayer networks. Scientific reports 6, 1–8 (2016).

26. Li, A., Cornelius, S. P., Liu, Y.-Y., Wang, L. & Barabási, A.-L. The fundamental advantages of temporal networks. Science 358, 1042–1046 (2017).

27. Baggio, G., Bassett, D. S. & Pasqualetti, F. Data-driven control of complex networks. Nature communications 12, 1–13 (2021).

28. Liu, Y.-Y. & Barabási, A.-L. Control principles of complex systems. Reviews of Modern Physics 88, 035006 (2016).

29. Löber, J. Optimal trajectory tracking of nonlinear dynamical systems (Springer, 2016).

30. Lai, S. et al. Effect of non-pharmaceutical interventions to contain covid-19 in china. Nature 585, 410–413 (2020).

31. López, L. & Rodó, X. The end of social confinement and covid-19 re-emergence risk. Nature Human Behaviour 4, 746–755 (2020).

32. COVID, T. I., Reiner, R., Barber, R. & Collins, J. Modeling covid-19 scenarios for the united states. Nature medicine (2020).

33. Keeling, M. J. & Rohani, P. Modeling infectious diseases in humans and animals (Princeton University Press, 2011).

34. Thompson, M. G. et al. Interim estimates of vaccine effectiveness of bnt162b2 and mrna-1273 covid-19 vaccines in preventing sars-cov-2 infection among health care personnel, first responders, and other essential and frontline workers (2021).

35. Stewart, G., Heusden, K. & Dumont, G. How control theory can help us control covid-19. IEEE Spectrum 57, 22–29 (2020).

36. Gertler, P. J., Martinez, S., Premand, P., Rawlings, L. B. & Vermeersch, C. M. Impact evaluation in practice (The World Bank, 2016).

37. Dowdle, W. R. The principles of disease elimination and eradication. Bulletin of the World Health Organization 76, 22 (1998).

38. Wang, R. et al. Analysis of sars-cov-2 mutations in the united states suggests presence of four substrains and novel variants. Communications Biology 4, 1–14 (2021).

39. Chowdhury, M. A., Hossain, N., Kashem, M. A., Shahid, M. A. & Alam, A. Immune response in covid-19: A review. Journal of Infection and Public Health (2020).

40. Lee, A., Thornley, S., Morris, A. J. & Sundborn, G. Should countries aim for elimination in the covid-19 pandemic? bmj 370 (2020).

41. Heywood, A. E. & Macintyre, C. R. Elimination of covid-19: what would it look like and is it possible? The Lancet Infectious Diseases 20, 1005–1007 (2020).

42. Metcalf, C. J. E., Morris, D. H. & Park, S. W. Mathematical models to guide pandemic response. Science 369, 368–369 (2020).

43. Thompson, R. N. et al. Key questions for modelling covid-19 exit strategies. Proceedings of the Royal Society B 287, 20201405 (2020).

44. Covid-19 vaccinations in the united states. https://covid.cdc.gov/covid-data-tracker/vaccinations. Accessed: 2021-03-2.

45. Baker, M. G., Kvalsvig, A., Verrall, A. J., Telfar-Barnard, L. & Wilson, N. New zealand’s elimination strategy for the covid-19 pandemic and what is required to make it work. The New Zealand Medical Journal (Online) 133, 10–14 (2020).

46. Diekmann, O., Heesterbeek, H. & Britton, T. Mathematical tools for understanding infectious disease dynamics, vol. 7 (Princeton University Press, 2012).

47. Oldenburg, B. & Glanz, K. Diffusion of innovations. Health behavior and health education: Theory, research, and practice 4, 313–333 (2008).

48. Flaxman, S. et al. Estimating the effects of non-pharmaceutical interventions on covid-19 in europe. Nature 584, 257–261 (2020).

49. Group, W. H. O. W. Nonpharmaceutical interventions for pandemic influenza, national and community measures. Emerging infectious diseases 12, 88 (2006).

50. Qi, J., Wang, S., Fang, J.-a. & Diagne, M. Control of multi-agent systems with input delay via pde-based method. Automatica 106, 91–100 (2019).

51. Smyshlyaev, A. & Krstic, M. Adaptive control of parabolic PDEs (Princeton University Press, 2010).

52. Agachi, P. S., Cristea, M. V., Csavdari, A. A. & Szilagyi, B. Model predictive control (De Gruyter, 2016).

53. De Silva, C. W. Intelligent control: fuzzy logic applications (CRC press, 1995).

54. Dong, E., Du, H. & Gardner, L. An interactive web-based dashboard to track covid-19 in real time. The Lancet infectious diseases 20, 533–534 (2020).

55. Safegraph. https://covid.cdc.gov/covid-data-tracker/national-lab. Accessed: 03 20 2021.

56. Covid-19 public monitor. https://today.yougov.com/covid-19. Accessed: 03 20 2021.

57. Gao, F. & Han, L. Implementing the nelder-mead simplex algorithm with adaptive parameters. Computational Optimization and Applications 51, 259–277 (2012).

