## Supplementary file for "Vaccination and three non-pharmaceutical interventions determine the end of COVID-19 at 381 metropolitan statistical areas in the US"

May 4, 2021

#### Contents

**1 Controllability of the SIRD Model** **2**

**2 Selection of NPIs for parsimonious model** **3**

**3 Robustness of feedback control system** **4**

**4 Vaccination Rollout** **5**

### 1 Controllability of the SIRD Model

For the nonlinear dynamical system,

$$\dot{X} = F(X, U), \tag{S1}$$

where $\dot{X} = [\dot{S}, \dot{I}, \dot{R}, \dot{D}]^T$ is the vector/output trajectory and $U = U_\Theta + \Theta_0$ is the inputs. Assume that the motion of the nonlinear system is in the neighborhood of the nominal system trajectory, that is

$$X(t) = X_n + \Delta X(t) \tag{S2}$$

where $\Delta X(t)$ represents a small quantity. Then,

$$U(t) = U_n + \Delta U(t) \tag{S3}$$

Thus, we have

$$\dot{X}_n(t) + \Delta \dot{X}(t) = F(X_n + \Delta X(t), U_n + \Delta U(t)) \tag{S4}$$

with $X_n = [S_n, I_n, R_n, D_n]^T$ and $U_n = [\beta_n, \gamma_n, \delta_n]^T$. Since $\Delta X(t)$ and $\Delta U(t)$ are small quantities, the right-hand side can be expanded into a Taylor series about the system trajectory and input, which produces

$$\dot{X}_n(t) + \Delta \dot{X}(t) = F(X_n, U_n) + \frac{\partial F}{\partial X}(X_n, U_n)\Delta X(t) + \frac{\partial F}{\partial U}(X_n, U_n)\Delta U(t) \tag{S5}$$

where higher orders are abandoned. The linear differential equation is obtained as

$$\Delta \dot{X}(t) = \frac{\partial F}{\partial X}(X_n, U_n)\Delta X(t) + \frac{\partial F}{\partial U}(X_n, U_n)\Delta U(t) \tag{S6}$$

with $\Delta X(t) = [\Delta S(t), \Delta I(t), \Delta R(t), \Delta D(t)]^T$, $\Delta U(t) = [\Delta U_\beta(t), \Delta U_\gamma(t), \Delta U_\delta(t)]^T$. Then,

$$\Delta \dot{X}(t) = A\Delta X(t) + B\Delta U(t) \tag{S7}$$

with $A = \frac{\partial F}{\partial X}(X_n, U_n)$

$$A = \begin{bmatrix} -\beta_n I_n & \mu - \beta_n S_n & \mu & 0 \\ \beta_n I_n & \beta_n S_n - \gamma_n - \delta_n - \mu & 0 & 0 \\ 0 & \gamma_n & \mu & 0 \\ 0 & \delta_n & 0 & 0 \end{bmatrix} \tag{S8}$$

and $B = \frac{\partial F}{\partial U}(X_n, U_n)$,

$$B = \begin{bmatrix} -S_n I_n & 0 & 0 \\ S_n I_n & -I_n & -I_n \\ 0 & I_n & 0 \\ 0 & 0 & -I_n \end{bmatrix} \tag{S9}$$

Then that system of Eq. S1 is controllable needs the full rank of following matrix.

$$C = [B \vdots AB \vdots ... \vdots A^{n-1}B] = \begin{bmatrix} -S_n I_n & 0 & 0 & \vdots & ...... \\ S_n I_n & -I_n & 0 & \vdots & ...... \\ 0 & I_n & 0 & \vdots & ...... \\ 0 & 0 & -I_n & \vdots & ...... \end{bmatrix} \tag{S10}$$

Since the $rank(C) = 4$, the system of Eq. S1 is controllable.

### 2 Selection of NPIs for parsimonious model

It's known that the low parsimony model (adding more parameters in the model) tends to have a better fit than the high parsimony model. To get a model with the goodness of fit and high parsimony, we test the human behaviour adherence to multiple NPIs data in Tab.S1 and Fig. S1. The NPIs range from the stay-at-home order, face-mask wearing, and testing capacity in the manuscript to the quarantine, school closure, and personal measures to avoid COVID-19

Suppose multiple NPIs $\boldsymbol{\theta}_I = [\theta_1, \theta_2, ..., \theta_N]^T$, we divide them into two sets, i.e., the set of community NPIs $\vartheta^c$ and the set personal NPIs $\vartheta^p$, see the type Tab. S1. Then the potential parsimonious model is

$$\begin{aligned} \hat{U}_\Theta(t) &= f(\boldsymbol{\theta}_I(t), \boldsymbol{W}_\Theta^I) \\ &= \prod_{i \in \{c,p\}} (1 - \sum_{j \in \vartheta^i} w_\Theta^j \theta_j(t)) - 1 \end{aligned} \tag{S11}$$

Thus, we test the accuracy of potential models against the different combinations of NPIs. The accuracy is measured by the mean absolute error between $\hat{U}_\Theta$ and the designed $U_\Theta$ derived from the feedback controllers at each MSA $m$ at times $t$ as

$$\begin{aligned} r_\beta &= \frac{1}{T} \sum_m \sum_t |\hat{U}_\beta^m(t) - U_\beta^m(t)| \\ r_\gamma &= \frac{1}{T} \sum_m \sum_t |\hat{U}_\gamma^m(t) - U_\gamma^m(t)| \\ r_\delta &= \frac{1}{T} \sum_m \sum_t |\hat{U}_\delta^m(t) - U_\delta^m(t)| \end{aligned} \tag{S12}$$

The results in Fig. S1b-d shows that more NPIs are included in Eq. S11 usually lead to higher accuracy with lower value of $r_\beta$, $r_\gamma$, and $r_\delta$. However, the accuracy comes to the highest around three or four NPIs are included. Tab.

S2 shows that the combination of the three NPIs, i.e., stay-at-home order, face-mask wearing, and testing enable the above model with high parsimony and better fitness with designed controllers.

### 3 Robustness of feedback control system

To extensively validate the SIRD-based feedback control system, we test its robustness in predicting local infections for (1) MSAs with geographical diversities; (2) seroprevalence estimates, including the estimated under-reporting cases with mild illness or no symptoms.

**All MSAs.** In the manuscript, we have shown the high predictive accuracy of the system for different MSAs. Here, we provide additional supportive results for the argument. Firstly, the average of MSAs' proportions of the infection rate over recover rate keeps constant around 2.664, as shown in Fig. S2. In contrast, the average of MSAs' proportions of the infection rate over death rate keeps increasing. It indicates, for simplicity, considering the NPIs' effects on $U_\beta(t)$ and $U_\delta(t)$ is enough to retrieve the input vector $U = [U_\beta, U_\gamma, U_\delta]^T$ for $U_\gamma = U_\beta/2.664$. Secondly, we used the R-squared value ($R^2$) to quantify daily predictive accuracy between all MSAs' predicted infection/recovery/death cases and their reported infection/recovery/death cases. From 1 April 2020 to 20 February 2021, the $R^2$ persistently greater than 0.9. The results indicate the robustness of the feedback control system over MSAs' geographical diversities. Thirdly, though the stay-at-home order and facemask wearing mainly negatively affect MSAs, the testing varies. Fig. S4 shows the needed magnitudes for the three NPIs (i.e., stay-at-home order, facemask wearing, and testing) at the 12 most populous MSAs in the US. For MSAs having positive effects for testing intervention, the needed magnitudes for stay-at-home order and facemask wearing become larger as testing gets larger. On the contrary, see the examples of ”Los Angles” MSA and ”Miami” MSA having negative effects for testing intervention. The needed magnitudes for stay-at-home order and facemask wearing become smaller as testing gets larger.

**Seroprevalence Estimates.** The Nationwide Commercial Laboratory Seroprevalence Survey develops large-scale geographic seroprevalence studies to learn more about the percentage of people in the US who have been infected with COVID-19 by states [1, 2]. By taking the average of states' seroprevalence estimates at each MSA and divide it by the reported infection data, we got each MSA's under-reporting ratios. Keeping the death cases the same, we obtain the estimated infected/recovered cases with under-reporting ratios. Fig. S5a shows the MSA's under-reporting ratios from 14 February 2020 to 20 February 2020. The MSAs' under-reporting ratios are relatively homogeneous, around 2.5. Compared to the reported data, the controllers on infection rate $U_\beta$ is larger. In contrast, the controllers on recovery rate $U_\gamma$ and death rate $U_\delta$ are smaller as shown in Fig. S5b. The learned marginal effects learned from seroprevalence data are in small deviations from the reported data. As shown in Fig. S5b-c, except for $w_\delta^s$, most of MSAs' effects of $w_\beta^s$, $w_\beta^f$, $w_\beta^g$, $w_\delta^f$, and $w_\delta^g$ are in the diagonal lines. These results demonstrate that the feedback control system has the predictability for the seroprevalence estimate data.

### 4 Vaccination Rollout

Fig. S6 shows how different MSAs gradually diminish the infection with different vaccination coverage locally. Though the elimination of COVID-19 would be a slow process that would have a long ”tail” of newly infected cases with random fluctuations, it could not be observed in our experiment as we didn't consider the spatial heterogeneity and reimportation from other areas. All the experiments indicate the ”New York” MSA has the highest new infected cases among all the MSAs, meaning it needs more rigorous NPIs and mass vaccinations to lower the possible infection and death in the future. As shown in Fig. S7, though the elimination days are relatively homogenous across MSAs, the additional dead cases are more heterogenous across MSAs. ”New York” MSA and 'Los angels' MSA have far more deadly than other MSAs, followed by ”Chicago” MSA, ”Dallas” MSA, ”Phoenix” MSA, ”Boston” MSA, and ”Philadelphia” MSA.

Table S1: Non-pharmaceutical interventions considered in this study

| NPIs | Explanation | Type |
|---|---|---|
| stay-at-home order | the normalized ratio of excessive time of staying at home | community NPI |
| school closure | fraction of people support for shool closure | community NPI |
| quarantine | fraction of people willing to be quarantined after contacting infected people | community NPI |
| working from home | fraction of people willing working from home | community NPI |
| face-mask wearing | the fraction of people wearing face masks | personal NPI |
| testing | fraction of tested population | personal NPI |
| frequent hand wash | fraction of people improve personal hygiene | personal NPI |
| Avoid crowding | fraction of people avoiding crowded places | personal NPI |

Table S2: Ranking of combinations of three non-pharmaceutical interventions having higher accuracy (i.e., lower deviation from designed controllers).

| rank | combinations of NPIs | $r_\beta$ | $r_\gamma$ | $r_\delta$ |
|---|---|---|---|---|
| 1 | {stay-at-home order, face-mask wearing, testing } | 10.311 | 3.078 | 0.568 |
| 2 | {stay-at-home order, face-mask wearing, avoid crowding } | 10.344 | 3.058 | 0.513 |
| 3 | {stay-at-home order, face-mask wearing, work from home } | 10.429 | 3.206 | 0.582 |
| 4 | {quarantine, face-mask wearing, work from home } | 10.473 | 3.116 | 0.538 |
| 5 | {frequent hand wash ,face-mask wearing, work from home} | 10.642 | 3.204 | 0.571 |

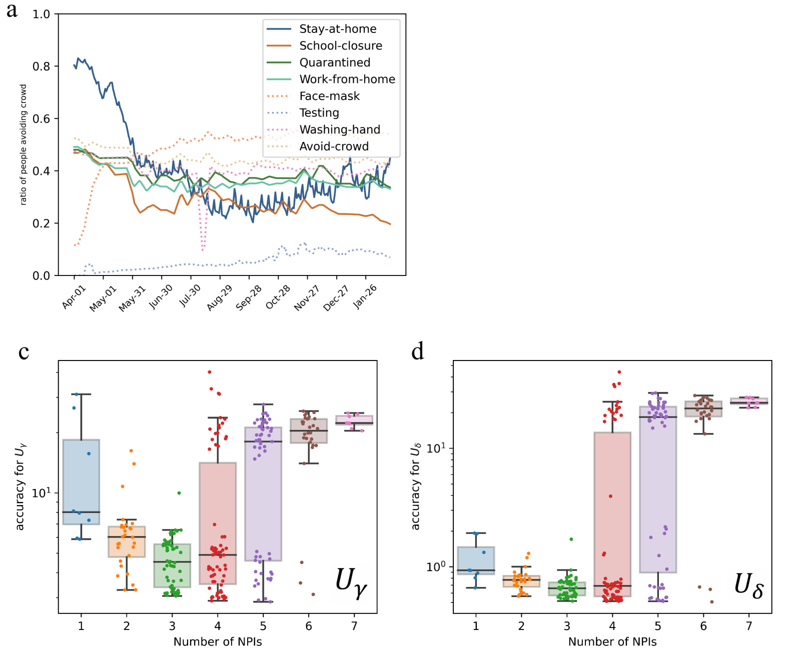


Figure S1: **Model parsimony versus goodness of fit. a** Temporal human adherence toward NPIs in Tab. S1. **b,c,d** The goodness of fit for controllers (i.e., $U_\beta$, $U_\gamma$, $U_\delta$) of the model Eq. S11 in against of number of NPIs included. Combined this figure with Tab. S2, it's easy to find that the stay-at-home order, face-mask wearing, and testing are most representative with higher accuracy in fitting controllers.

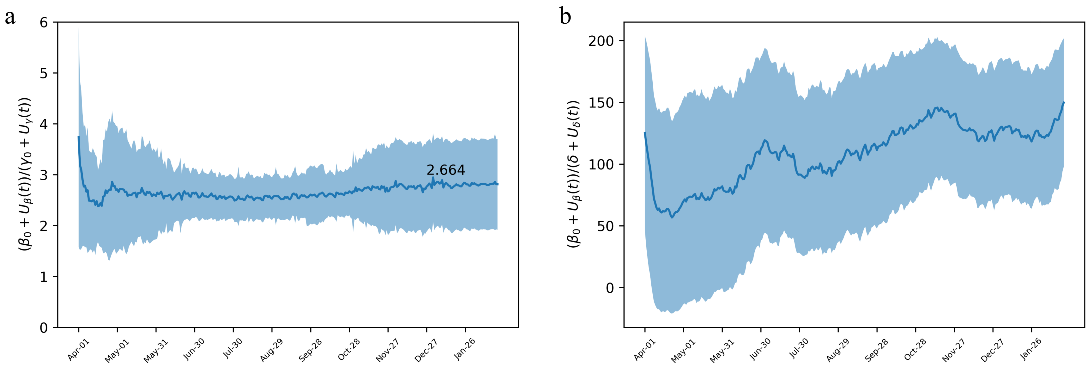


Figure S2: **Proportion of infection rate over recovery rate, and proportion of infection rate over death rate.** The solid line is the average proportions of all MSAs and the shadowed area is the variances of proportions.

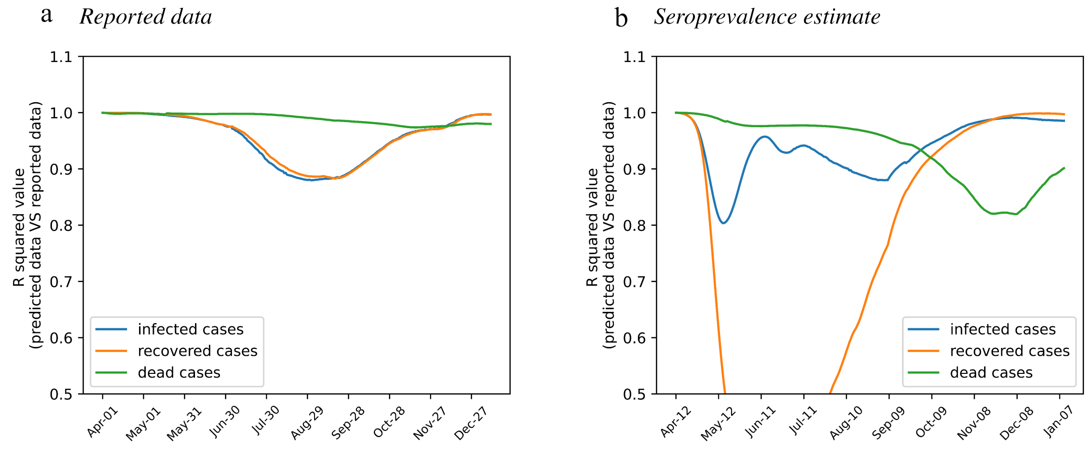


Figure S3: **R-squared values** for **a** measuring the accuracy of the predicted infection/recovery/death data with the reported data and **b** measuring the accuracy of the predicted infection/recovery/death data with the seroprevalence estimated data.

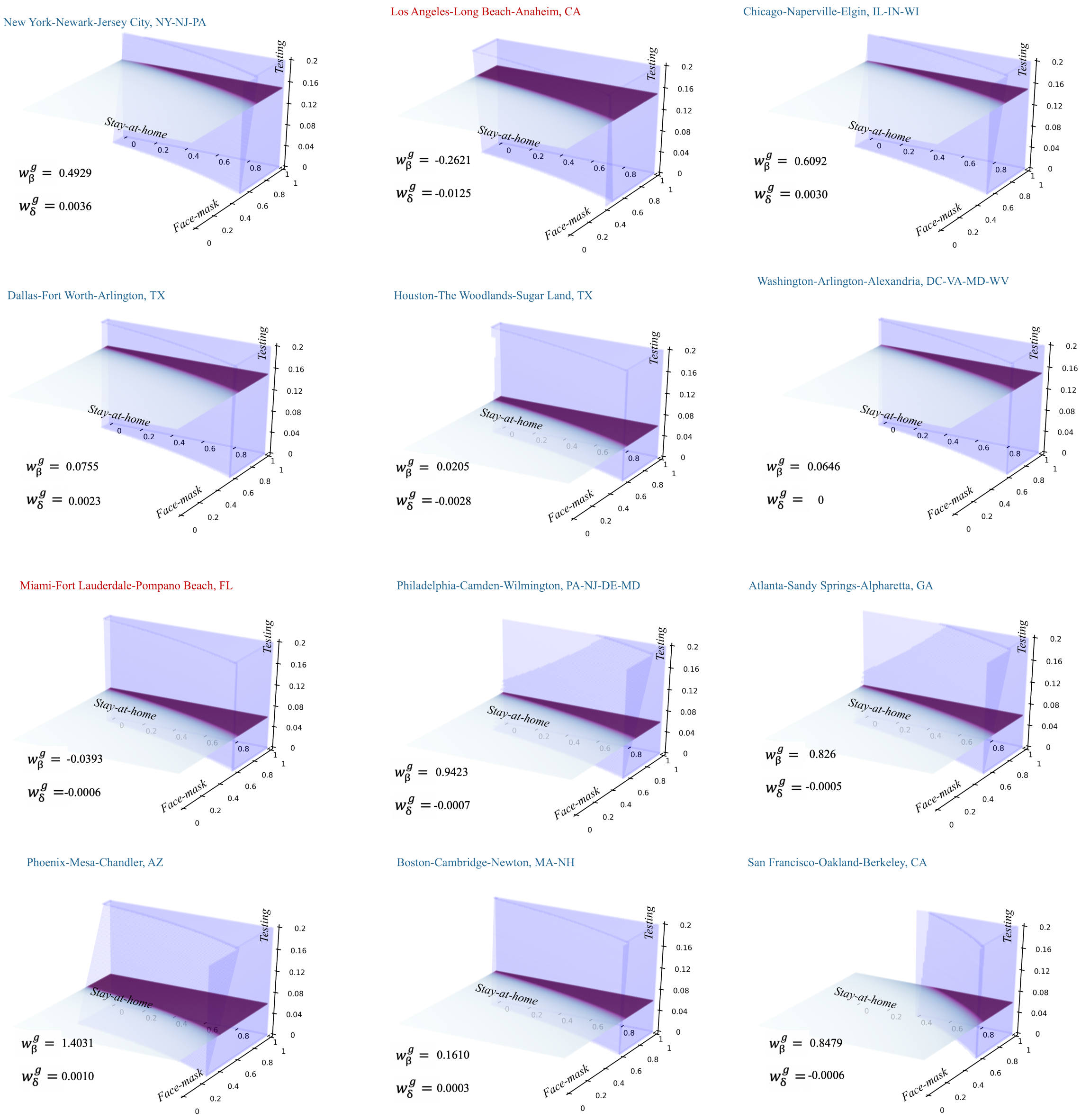


Figure S4: Needed magnitude for NPIs in order to keep $R_e \leq 1$ with zero vaccine coverage. The horizontal slices of the "triangular prism" are the visualization for the needed magnitude of stay-at-home order and face-mask wearing if the magnitude of testing is fixed as each MSA' recent testing capacity. The MSAs ("Los angels" MSA and "Miami" MSA) which are marked in red are those having negative effects for testing intervention. Different with other MSAs, the needed stay-at-home order and face-mask wearing gets smaller in "Los angels" MSA and "Miami" MSA as their testing capacities increase.

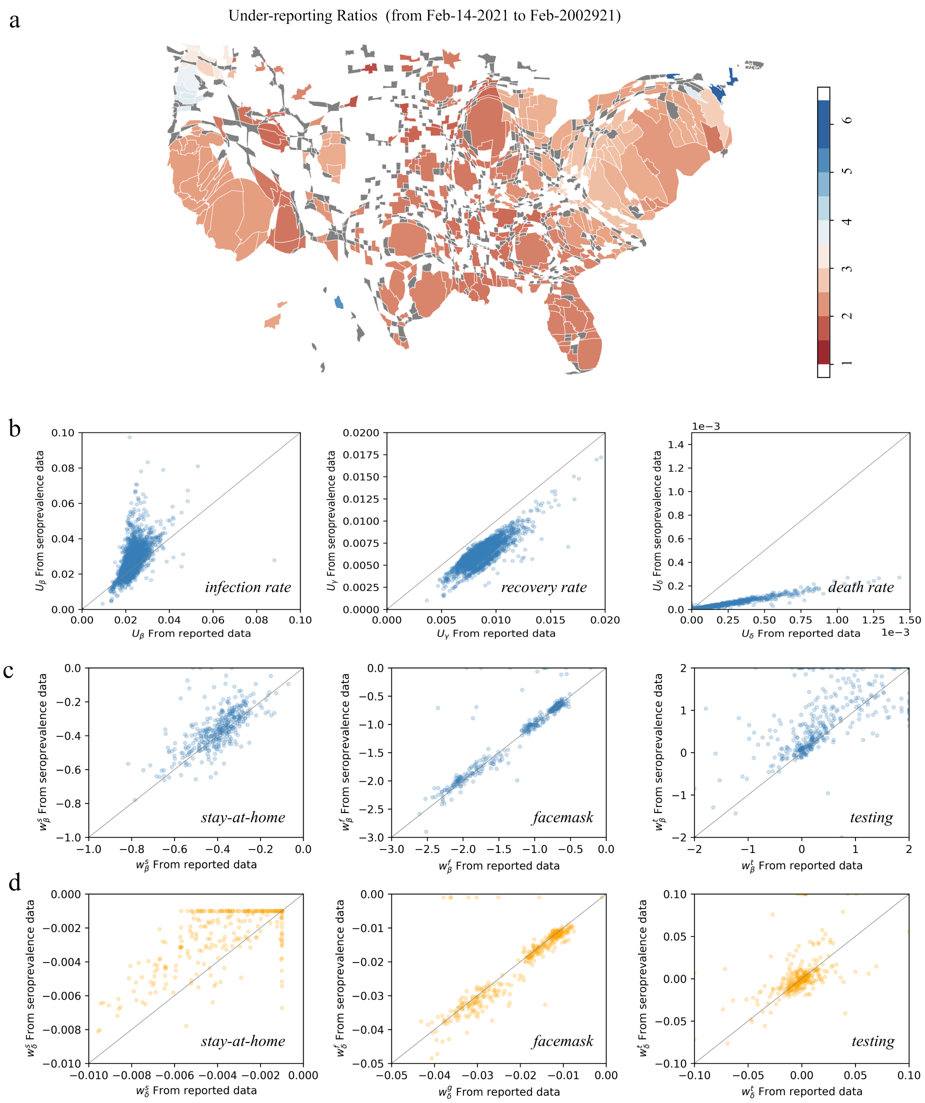


Figure S5: **Comparison of feedback controllers and learned NPIs' marginal effects when using the seroprevalence estimates and when using the reported infection data.** **a** Average under-reporting ratios (i.e., seroprevalence estimates over reported data) at MSAs from 14 February 2021 to 20 February 2021. **b** comparisons of controllers ($U_\beta$, $U_\gamma$, and $U_\delta$). **c,d** Comparisons of NPIs' marginal effects for $U_\beta$ and $U_\delta$.

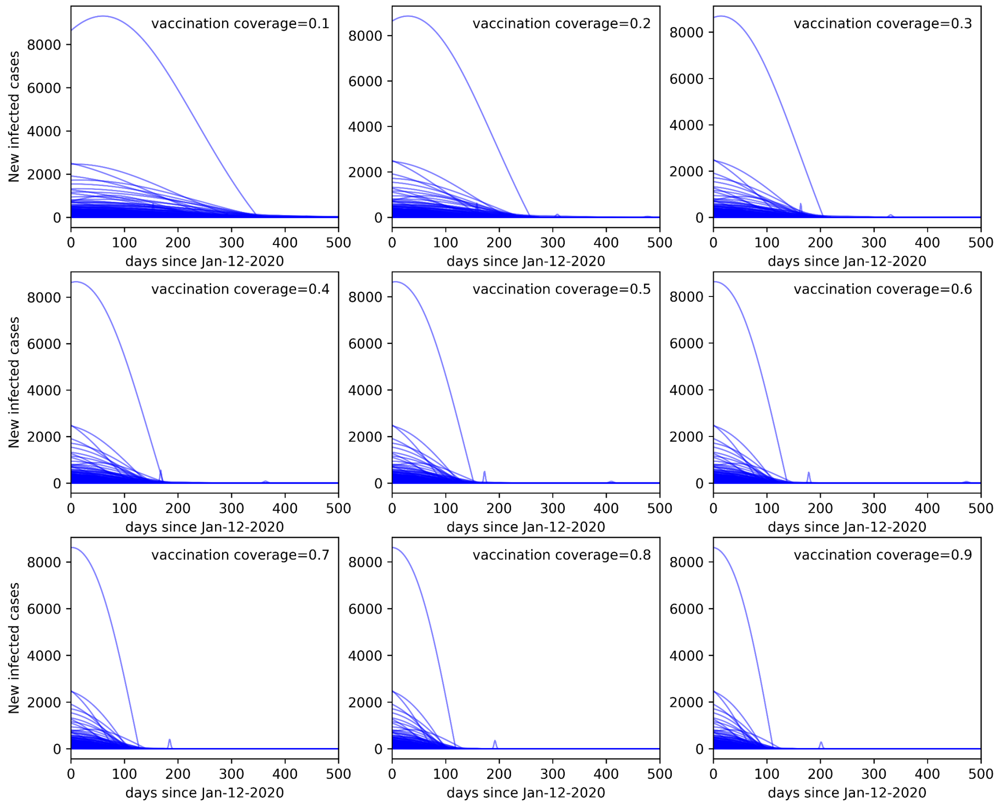


Figure S6: **Daily new infected cases since 12 January 2020 for different vaccination coverage within 360 days.**

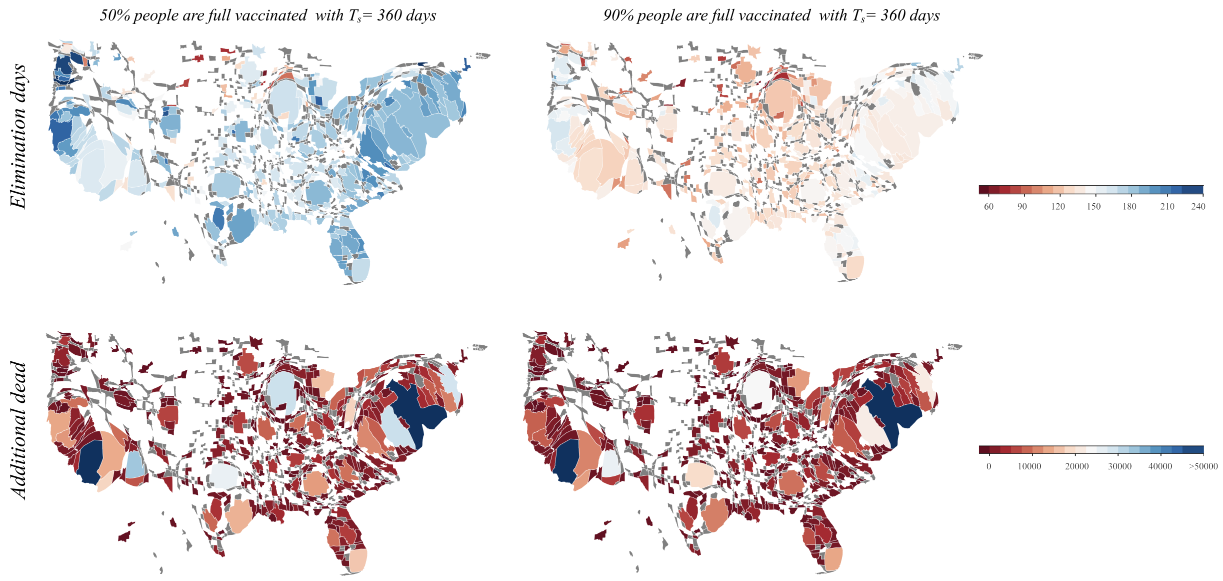


Figure S7: **MSAs' elimination days and additional dead cases if 50% or 90% people are full vaccinated in 360 days.**

### References

[1] Bajema, K. L. *et al.* Estimated sars-cov-2 seroprevalence in the us as of september 2020. *JAMA internal medicine* (2020).

[2] Nationwide commercial laboratory seroprevalence survey. https://www.safegraph.com/covid-19-data-consortium. Accessed: 2021-03-20.
